# High Resolution Multi-delay Arterial Spin Labeling with Transformer based Denoising for Pediatric Perfusion MRI

**DOI:** 10.1101/2024.03.04.24303727

**Authors:** Qinyang Shou, Chenyang Zhao, Xingfeng Shao, Megan M Herting, Danny JJ Wang

**Author notes:** (Corresponding Author: Danny JJ Wang).

## Abstract

Multi-delay arterial spin labeling (MDASL) can quantitatively measure cerebral blood flow (CBF) and arterial transit time (ATT), which is particularly suitable for pediatric perfusion imaging. Here we present a high resolution (iso-2mm) MDASL protocol and performed test-retest scans on 21 typically developing children aged 8 to 17 years. We further proposed a Transformer-based deep learning (DL) model with k-space weighted image average (KWIA) denoised images as reference for training the model. The performance of the model was evaluated by the SNR of perfusion images, as well as the SNR, bias and repeatability of the fitted CBF and ATT maps. The proposed method was compared to several benchmark methods including KWIA, joint denoising and reconstruction with total generalized variation (TGV) regularization, as well as directly applying a pretrained Transformer model on a larger dataset. The results show that the proposed Transformer model with KWIA reference can effectively denoise multi-delay ASL images, not only improving the SNR for perfusion images of each delay, but also improving the SNR for the fitted CBF and ATT maps. The proposed method also improved test-retest repeatability of whole-brain perfusion measurements. This may facilitate the use of MDASL in neurodevelopmental studies to characterize typical and aberrant brain development.

## I. INTRODUCTION

The developmental period between childhood to adolescence and young adulthood is associated with the emergence of a number of behavioral and emotional disorders that may last lifelong. It is critical to understand detailed patterns of typical development, so alterations can be identified and rectified as early as possible. As an entirely noninvasive and quantitative imaging method, arterial spin labeled (ASL) perfusion MRI is increasingly being recognized as an important biomarker for functional brain development in both healthy populations and neurodevelopmental disorders[1]. However, there remain significant challenges for making ASL an impactful tool in studying neurodevelopment, including: 1) Existing ASL techniques generally have a coarse spatial resolution of ∼4×4×4mm^3^, making it difficult to decompose structural and functional components of developmental effects due to partial volume effects; and 2) The recommended segmented 3D acquisition[2] takes relatively long scan time and generally allows a single post-labeling delay (PLD) scan, which is susceptible to age dependent variations in arterial transit time (ATT), affecting the accuracy of perfusion quantification. Multi-delay ASL can provide improved accuracy for quantitative measurement of both CBF and ATT compared to single-delay ASL[3]. However, it needs to divide the total scan time to acquire multiple delays and thus sacrificing SNR of each delay, which makes acquiring high-resolution multi-delay ASL images even more challenging.

In order to achieve robust high-resolution multi-delay ASL, accelerated image acquisition/reconstruction and denoising techniques are desired, which may be applied separately or jointly through denoising-reconstruction techniques. Compressed Sensing [4] is a widely used technique, which uses L1 wavelet or total variation (TV) regularizer for denoising and reconstruction. In another previous study[5], the authors proposed a robust single-shot acquisition of multi-delay ASL with time dependent CAIPIRINHA under-sampling pattern and spatiotemporal reconstruction with total generalized variation (TGV) regularization. However, these methods require predefined constraints for optimization, and generally take a relatively long time for image reconstruction due to iterative optimization steps. Another way to denoise multi-delay ASL images is to use k-space weighted image average (KWIA)[6], which divides the k-space into multiple rings and applies a progressively wider temporal window for moving average of peripheral k-space data to reduce noise. This method does not require predefined constraints or iterations, and preserves the spatiotemporal resolution of the original image series. However, the image noise becomes temporally correlated following KWIA denoising, resulting in little improvement on the fitting results, i.e., CBF and ATT maps[6].

Deep learning (DL) provides a powerful tool for medical image processing[7]. DL has been applied for ASL denoising [8], [9], [10], [11] with either CNN or Transformer-based networks. DL methods can process noisy images efficiently and effectively once the model has been trained. However, training a DL model usually relies on paired images with a low-SNR image as input and a high-SNR image as the reference. This makes it hard to apply to cases where there are limited or no high-SNR images acquired, especially for a high-resolution dataset where acquiring such a reference image is very time-consuming. While using an existing pretrained model may in part help with this issue, the performance of the model may decline on a new dataset with different data distribution such as different image contrast or different image resolution, and usually fine-tuning of the pretrained model is warranted.

In this work, we first developed a high-resolution (iso-2mm) multi-delay ASL imaging protocol for pediatric perfusion imaging. We further proposed a DL denoising framework for multi-delay ASL with KWIA denoised images as reference. We trained and tested the performance of the proposed DL model on a cohort of 21 typically developing children aged 8 to 17 years-old with test and retest scans. The proposed method was compared with several benchmark methods including TGV regularized reconstruction, KWIA and a pretrained Swin Transformer.

The main contributions of this work are summarized as following:

- We developed a high-resolution multi-delay ASL protocol for pediatric perfusion imaging, which was applied in children with test-retest scans to evaluate the performance of different methods;
- We proposed a novel DL framework for multi-delay ASL denoising where there are no acquired reference images to train the model;
- We used a CNN and Swin Transformer combined network in our network to improve the performance of denoising;
- Our proposed method was compared with state-of-the art methods including non-DL methods (KWIA, TGV) and different DL strategies (train-from-scratch, fine-tune and direct apply).

## II. Related work

### A. ASL acquisition with CAIPIRINHA acceleration and TGV reconstruction

The recommended acquisition for ASL according to the consensus paper[2] is to use a segmented 3D readout. In a recent study[5], the authors used a modified segmented 3D GRASE readout with a time dependent 2D CAIPIRINHA[12] pattern to improve SNR and robustness for acquiring multi-delay pseudo-continuous ASL (pCASL) images with iso-3mm resolution. In this work, we expanded the previous work with a higher acceleration factor to achieve high resolution (iso-2mm) multi-delay pCASL. The under-sampling pattern for each measurement is depicted in Figure 1. A total of 8 segments with an acceleration factor of 2 in the phase encoding (PE) (y) direction and 4 in the partition (z) direction were used in the readout to shorten the length of each echo train. There were shifts between subsequent control/label pair both in the PE and partition direction to increase temporal incoherence and reduce aliasing artifacts for reconstruction. There are several advantages for this pattern. First, it enables the estimation of coil sensitivity maps directly from the combined k-space without the need to acquire a separate calibration data, like regular GRAPPA-type reconstruction. Second, this pattern is more robust to motion artifacts compared to the standard segmented acquisition[5]. Third, it enables the options to reconstruct the image with either parallel imaging techniques for each segment or direct inverse Fourier Transform of the segment-combined k-space.

**Fig. 1.**
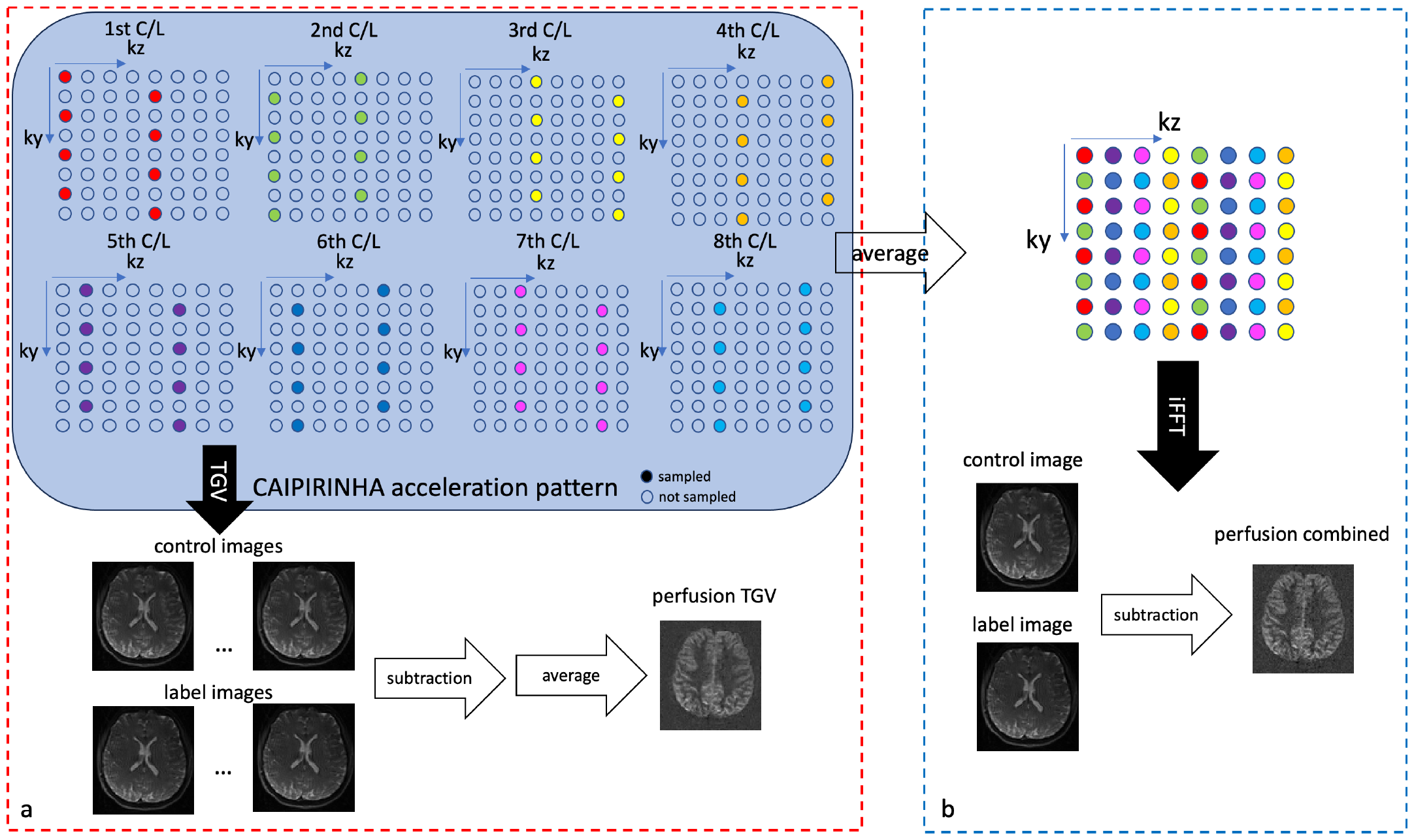
Illustration of the accelerated acquisition with CAIPI under-sampling pattern and two different approaches for reconstruction including combining the k-space and IFFT (a) and TGV constrained reconstruction for each k-space segment and average afterwards (b).

As the baseline processing pipeline for this type of under-sampled acquisition, a spatiotemporal TGV constrained reconstruction was applied to the k-space data for each PLD. The reconstruction solves the following optimization equation:

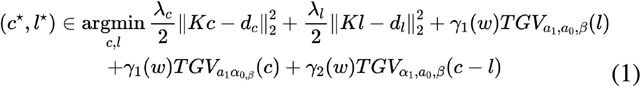

where *c* and *l* are control and label images for one PLD, *K* is the forward encoding matrix, including coil sensitivity profile, Fourier operation and under-sampling pattern masking, *d*_*c*_ and *d*_*l*_ are the acquired k space data for control and label respectively. *λ*_*c*_ and *λ*_*l*_ are weights to balance the data consistency term. α_0_, α_1_ and β are TGV reconstruction parameters according to[13] and γ_1_(w), γ_2_(w) are the weights to balance the regularization for control, label, and perfusion images. The TGV regularizer enforces piece-wise smooth images for both control/label and the perfusion image.

The TGV constrained reconstruction method can improve the SNR of the reconstructed perfusion images. However, the reconstruction is time-consuming and has high demand in GPU memory as it takes the k-space data with multiple coils and multiple time points into the reconstruction algorithm.

### B. KWIA denoising for multi-delay ASL

K-space weighted image average (KWIA) is a denoising technique originally proposed for computed tomography perfusion (CTP) imaging to reduce X-ray radiation dose [14] and later adapted to denoising dynamic MRI series such as MR angiography and multi-delay ASL[6].

KWIA performs denoising of an image within a time series by first dividing the whole k-space into several ring-shaped sub-k-space. The central k-space region uses original data to preserve the image contrast and temporal resolution, and the outer k-space is weighted averaged among neighboring time frames to increase SNR while preserving spatial resolution. The weighting for the averaged k-space is adjusted according to the number of neighboring time frames to be averaged. The KWIA processed k-space data can be described by the following equation:

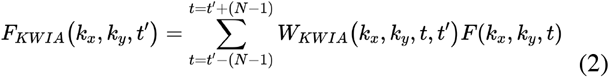

where *F*_*KWIA*_ and *F* denotes the k-space data after and before denoising respectively, and *W*_*KWIA*_ denotes the weight for the averaged rings. Figure 2 illustrates the processes of applying KWIA with 2 rings to 5-delay ASL data.

**Fig. 2.**
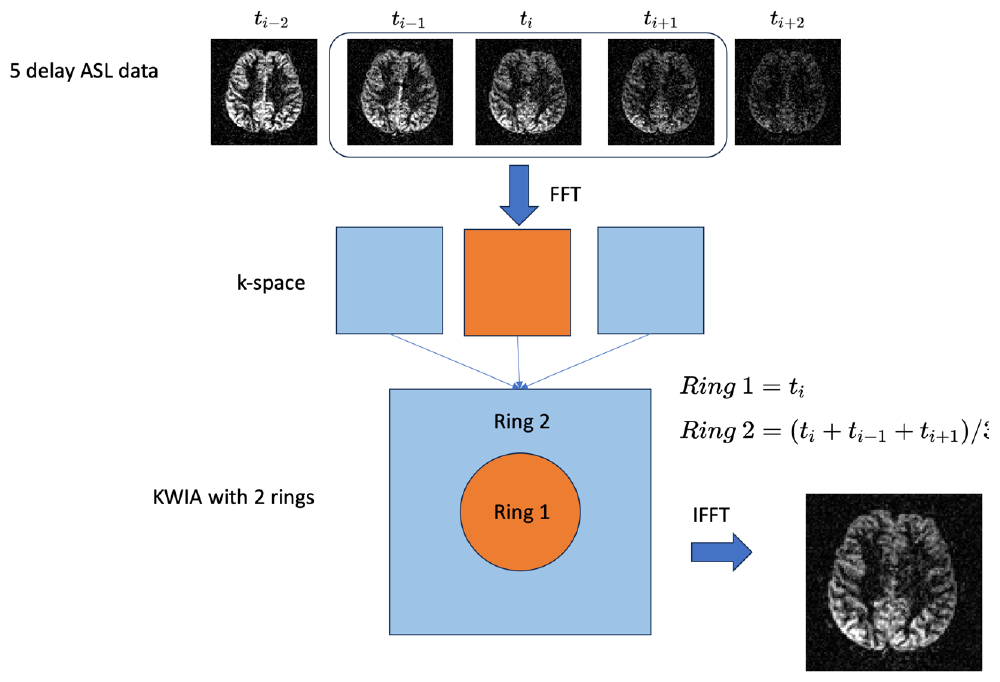
Illustration of the KWIA method with 2 rings to 5-delay ASL data.

The main idea of this method is based on that most of the image contrast and temporal information is stored in the low-frequency part of the k-space while the image resolution and fine details are determined by the extent of the k-space coverage. Therefore, averaging the outer rings while keeping the center k-space unchanged can improve the SNR while preserving the spatiotemporal resolution and contrast of the original image series.

The advantage of KWIA is that it can be easily implemented without complicated computation. It also does not cause much spatial blurring to the image like some traditional denoising methods such as Gaussian filtering. However, although theoretically KWIA can improve the SNR of each time frame image by about 2 folds, the results for quantitative fitting show little improvements since KWIA induces temporally correlated noise, thus does not help when all images are used for fitting[6]. As a result, KWIA can only improve the SNR and better delineation of image details for each time frame, but not improve the quantitative mapping.

### C. Deep Learning denoising for ASL

During the past decades, there have been several studies trying to denoise ASL images with deep learning methods. CNN-based networks with both regular and dilated convolutions have been developed to denoise for perfusion weighted images[8] or directly to the CBF maps[10]. A multi-contrast model that uses both M0 and perfusion images was used to further improve the denoising performance[9]. Recently, Swin Transformer-based networks that use window attention mechanism and window shifting techniques have been developed for image restoration tasks[15], [16] and was adapted to denoising both single and multi-delay ASL images[11] to beat the performance of the regular CNN-based networks.

These methods used a noisy image as the input and trained a model to produce an image with higher SNR. This is usually achieved by using a reference high-SNR image averaged with more repetitions than the input. In clinical settings, single-delay ASL is usually acquired with a relatively low-resolution and repeated multiple times to achieve a reasonable image quality. This averaged image can be used as the reference for denoising an image with fewer averages, thus reducing the scan time. However, since the SNR of MR images reduces with smaller voxel size, it would take longer scan time to achieve similar SNR for high-resolution acquisitions. Especially for multi-delay ASL, they are usually acquired with only a few or even only one repetition for each delay and therefore, no high-SNR images can be used as reference. In this case, the traditional supervised learning settings cannot be directly applied to train a model for denoising such high-resolution multi-delay ASL.

Given some desirable features of KWIA denoising including its computational efficiency and contrast preserving, and that it does not cause much spatial blurring, in this study, we proposed a Transformer based DL framework for denoising high-resolution multi-delay ASL using KWIA denoised images as reference. We show that the trained model can outperform KWIA in not only the SNR of the perfusion images, but also the SNR of the parametric maps (CBF and ATT).

### D. Pretrained model

Existing works of DL ASL denoising usually trained a new model on a specific dataset. However, in recent years, pretrained models have shown to be powerful especially in the case where there are not sufficient training data[17], [18], [19]. In general, a pretrained model is developed on a larger dataset with a more general task, which can be then applied to specific downstream tasks with fine-tuning. It has been shown that the pretrained model can provide a better starting point and need less time for training. But whether the performance of fine-tuning and training from scratch will differ is not clear, especially for a specific task such as ASL denoising.

In this study, besides training a new model from scratch, we also tried to utilize a pretrained model for lower resolution single-delay ASL denoising. We compared the performance of directly applying the pretrained model, training a new model from scratch and fine-tuning a pretrained denoising.

## III. Materials and methods

### A. Imaging Protocol and Datasets

In this study, we developed a high-resolution multi-delay ASL protocol with 8-fold 2D CAIPI acceleration. Twenty-one typically developing children (age=13±2.5 years, 13 males) without neurological/psychiatric disorders or development delays were recruited under IRB approval. All subjects were scanned twice with two weeks apart for test-retest purposes on a 3T MR system (Prisma, Siemens Healthcare, Germany) using a 32-channel head coil. The multi-delay ASL images were acquired with the proposed scheme, along with a separate T1w MPRAGE scan (1 mm^3^ isotropic) and a phase contrast MRA.

A time-dependent CAIPI under-sampling pattern with 8 segments was employed for each post labeling delay (PLD) for the multi-delay ASL protocol[5]. The complete imaging protocol is shown as follows: TR = 6180ms, TE = 52.5ms, FOV = 192×192×96 mm^3^, resolution of 2×2×2 mm^3^, matrix size of 96×96×48. A phase contrast MRA image was acquired before ASL and the labeling plane was placed at a straight segment of intracranial arteries and vertebral arteries to improve labeling efficiency. Labeling duration = 1500ms and 5 PLDs (600, 1000, 1400, 1800 and 2200ms) were used considering shorter ATT in younger populations[20]. Background suppression with 2 inversion pulses was used and optimized for each PLD[21]. The scan duration for this multi-delay ASL was 8 minutes and 33 seconds. This will result in a complete measurement (1 control and 1 label image) for each delay if averaging all segments, or 8 measurements if each segment is reconstructed respectively. Segment-combined images were used as the input for denoising. The above acquisition results in a dataset that contains a total of 42 scans with 5 delays and a total of 10080 slices.

### B. Network architecture

Both the pretrained model and the train-from-scratch model used a combined CNN and Swin Transformer[15], [16] network as the network architecture to perform ASL denoising. The network architecture is shown in Figure 3.

**Fig. 3.**
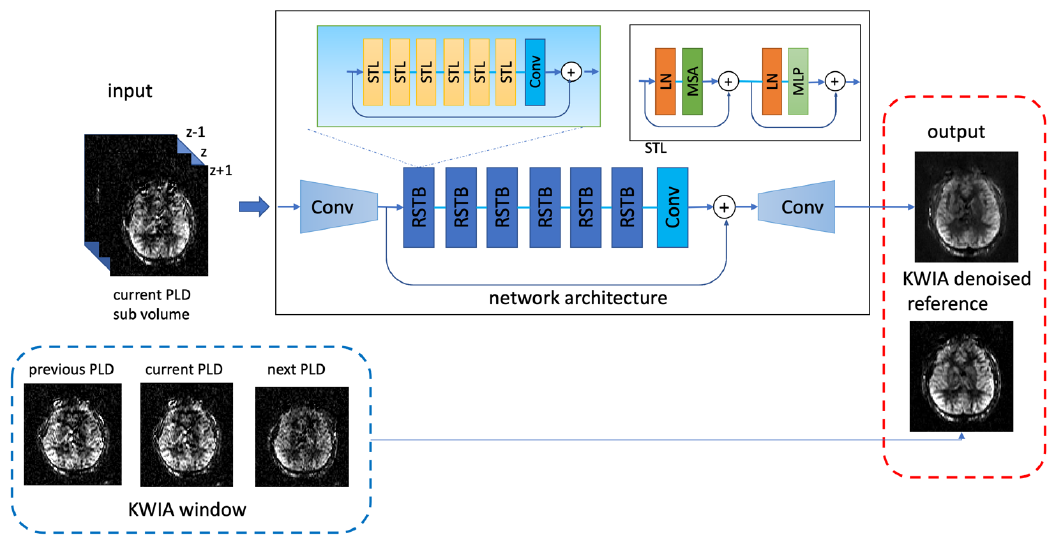
Illustration of the training framework and network structure.

The input layer takes the center and two adjacent slices of the image so the input size would be H×W×3. The first layer is a CNN layer that combines the pseudo3D input and extracts some shallow features of the image. The next few layers are Swin Transformer layers, which consist of several Swin Transformer blocks with a residual connection in each layer. The Swin Transformer layer is consisted of a multi-head self-attention and a two-layer multi-delay perceptron (MLP), with a layer normalization (LN) before each module and a residual connection. In two consecutive Swin Transformer layers, the first layer uses a regular windowed Transformer layer, and the second layer performs window shifting and takes the shifted windows into the Transformer operation. The computation of two consecutive Swin Transformer layers can be expressed as

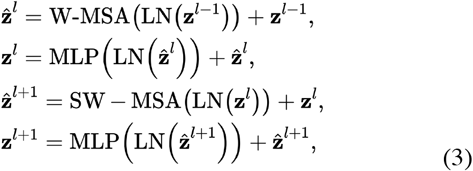

where 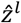 and *z*^*l*^ denote the outputs of the Shifted Window or regular MSA module and the MLP module for block *l* respectively. The last layer of the Swin Transformer block is a CNN layer, which brings the benefit of inductive bias of the convolution operation to improve the performance of regular Transformer operation.

The last layer of the whole network is a CNN layer to combine the features and produce the final denoised image. The output was 1-channel with the size of H×W and the loss function was computed with the output and the KWIA denoised image of that center slice.

### C. Implementation details

#### 1) TGV constrained reconstruction

TGV reconstruction algorithm was implemented in MATLAB with CUDA parallel processing using AVIONIC library[13]. Coil sensitivity maps were estimated using the segment-combined control images. The hyperparameters for the TGV optimization were chosen according to the preliminary experiments[5], where 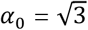, *α*_1_ = 1, w=0.9 and β=7. Regularization parameters *λ*_*c*_ = 10 and *λ*_*l*_ = 1.06 × *λ*_*c*_. For each PLD, 8 paired under-sampled k-space of control and label images were input to the algorithm and the output were 8 paired control, label and perfusion images. The average of the 8 output perfusion images were used as the final results for TGV reconstruction. (The reconstruction was run on a lambda cluster with NVIDIA RTX 3090 GPU and the algorithm took 126±1 seconds for each PLD.)

#### 2) KWIA denoising for multi-delay ASL

KWIA algorithm was implemented in MATLAB. A two-ring KWIA with the ratio of the centric circle to the radius of the entire k-space of 0.5 was used. Since the two-ring KWIA uses three time frames for denoising, for the first and last delay image, the same image was padded one frame to fulfill the requirement of KWIA. KWIA denoised images of each delay were used as the reference to train the DL model in the following steps.

#### 3) DL training

The network was implemented with Python 3.7 and Pytorch 1.12. Adam was used as the optimizer with the initial learning rate of 1e^-3^ and will decay if the performance in the validation data did not improve for 20 epochs. Batch size was 16 for all of the experiments. The loss function used in model training was a combination of L1 loss and structural similarity loss (SSIM loss).

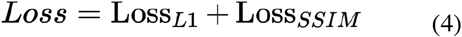

For training a new model (train from scratch), the model weights were randomly initialized. Fine-tuning uses the weights of the pretrained model as the starting point and all layers were updated during the training process. The pretrained model used the same network architecture as described in the previous session. It was trained on two single-delay ASL datasets with 104 training scans and has been tested on 57 scans from three different vendors. The details of the pretrained model was described in [11]. For comparison, we also tested the direct apply of the pretrained model to this new unseen dataset without retraining the model.

For both train-from-scratch and fine-tune model training, a five-fold cross validation was used which takes 27 cases for training, 8 cases for validation and 9 cases for testing. This allows that each case can be tested for once to evaluate the performance on the whole dataset.

### D. Results evaluation

To get the quantitative maps from the perfusion images, the original or the denoised multi-delay perfusion images were fed into the one compartment kinetic model[22] to fit for CBF and ATT maps using the following equation.

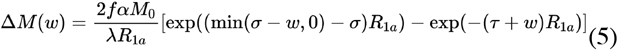

where f and σ are CBF and ATT to be fitted, ΔM (ω) is the measured perfusion signal, M0 is the equilibrium magnetization of brain tissue. Blood T1 and the corresponding relation rate R_1a_ was adjusted for each subject’s age and gender according to a previously established blood T1 model[23], α=0.78 is the labeling efficiency, λ =0.9 g/ml is the brain blood partition coefficient, τ=1.5s is the labeling duration and ω is PLD.

The performance of denoising was evaluated in three aspects. First, the quality of the denoised image is measured by gray matter SNR of the 1) perfusion images, 2) CBF maps and 3) ATT maps, calculated using the mean value in the gray matter (GM) divided by the standard deviation in the white matter (WM). Second, mean CBF and ATT values of whole brain, gray matter and white matter were calculated within the masks and compared to the values fitted from the original images without any denoising to assess the reconstruction accuracy. Third, intraclass correlation coefficient (ICC) of two-way mixed effects, absolute agreement and single rater[24] was calculated to evaluate the test and retest reliability from 21 subjects using the following equation:

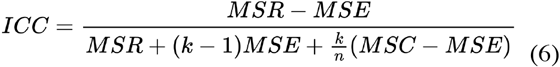

where MSR is the mean square for rows, MSE is the mean square for error, MSC is mean square for columns, n is the number of subjects and k is the number of measurements. ICC was calculated to evaluate the test-retest robustness of the acquisition and influence of the denoising algorithm.

## IV. EXPERIMENT RESULTS

### A. Denoising performance of perfusion images and quantitative maps

Figure 4 shows a representative case of the original and denoised perfusion images of 5 PLDs using traditional and proposed methods. Both non-DL and DL methods improved the SNR compared to the original segment-combined image. For all five delays, there were moderate improvements in SNR, even though SNR of the perfusion image of the longest PLD was lower than the shorter PLDs. The “direct apply” of the pretrained model yielded an oversmoothed results compared to other denoising methods, with less details in the subcortical areas.

**Fig. 4.**
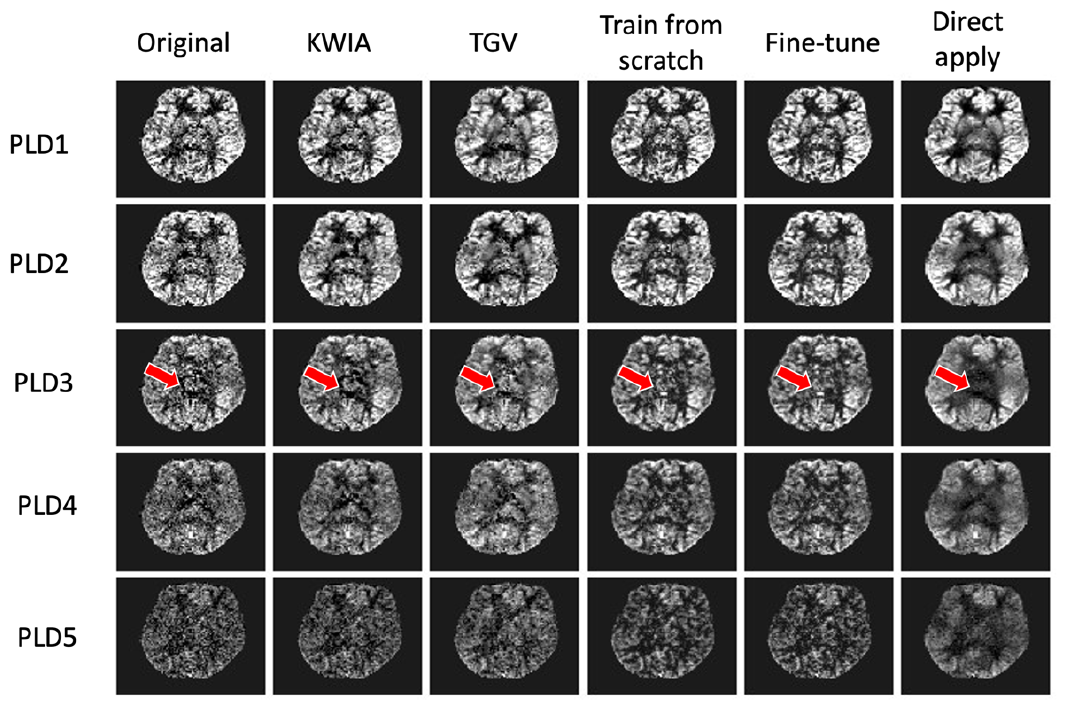
Original and denoised perfusion images of 5 delays. Red arrows show the subcortical areas.

Figure 5 shows a representative case of fitted CBF and ATT maps after applying different denoising methods. DL-based methods including both train-from-scratch and fine-tune show higher SNR in both CBF and ATT maps while also preserving the details in the gray and white matter (see zoom-in panels in Fig. 5).

**Fig. 5.**
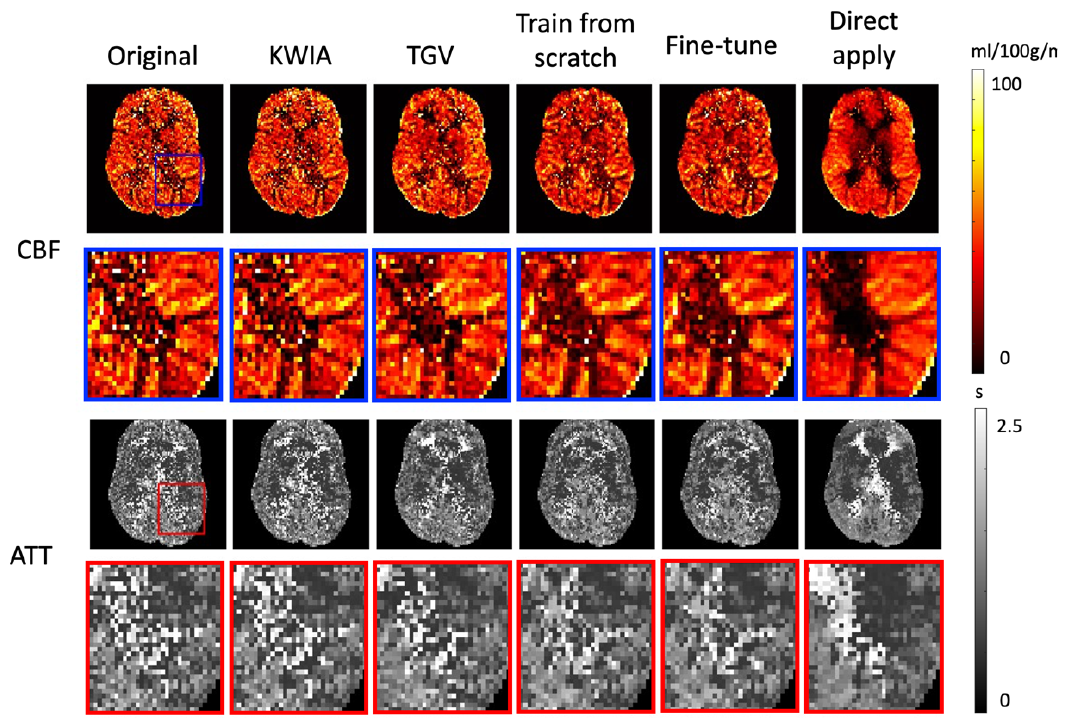
CBF and ATT maps of a representative subject processed with different denoising methods. The blue and red box show the selected area of zoom-in to visualize the details.

Figure 6 shows the bar plot of overall SNR performance of perfusion images, CBF and ATT maps on all cases. The performance of train-from-scratch model and fine-tune model are results averaged across 5 folds. All DL methods outperformed traditional methods including KWIA and TGV-based reconstruction across all 5 PLDs. For CBF and ATT maps, KWIA shows little improvements from the original, but DL-based methods trained on the KWIA-reference image show higher SNR than both KWIA and TGV method. The details of the results are shown in Table I and Table II.

**TABLE I.** SNR of perfusion images processed with different denoising methods.

| SNR | PLD1 | PLD2 | PLD3 | PLD4 | PLD5 |
| --- | --- | --- | --- | --- | --- |
| Original | 1.51±0.52 | 1.39±0.46 | 1.45±0.35 | 1.16±0.28 | 0.91±0.28 |
| KWIA | 1.79±0.62 | 1.86±0.62 | 1.95±0.49 | 1.61±0.39 | 1.11±0.34 |
| TGV | 2.24±0.78 | 2.14±0.72 | 2.17±0.59 | 1.68±0.46 | 1.22±0.37 |
| Train-from-scratch | 2.83±1.03 | 2.76±0.99 | 2.96±0.82 | 2.35±0.69 | 1.63±0.56 |
| Fine-tune | 2.79±1.06 | 2.76±1.06 | 2.90±0.88 | 2.28±0.66 | 1.60±0.54 |
| Direct apply | 3.98±1.38 | 3.69±1.32 | 4.01±1.14 | 3.15±0.90 | 2.18±0.81 |

**TABLE II.**
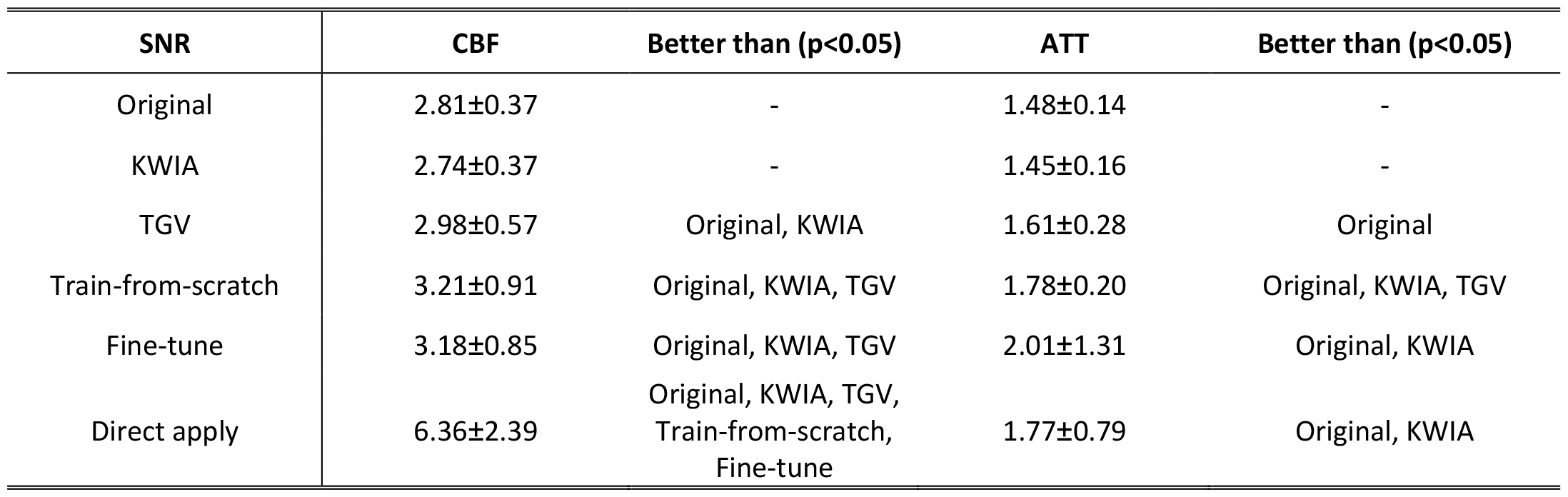
SNR of CBF and ATT maps with different denoising methods.

| SNR | CBF | Better than ($p<0.05$) | ATT | Better than ($p<0.05$) |
| --- | --- | --- | --- | --- |
| Original | 2.81±0.37 | - | 1.48±0.14 | - |
| KWIA | 2.74±0.37 | - | 1.45±0.16 | - |
| TGV | 2.98±0.57 | Original, KWIA | 1.61±0.28 | Original |
| Train-from-scratch | 3.21±0.91 | Original, KWIA, TGV | 1.78±0.20 | Original, KWIA, TGV |
| Fine-tune | 3.18±0.85 | Original, KWIA, TGV | 2.01±1.31 | Original, KWIA |
| Direct apply | 6.36±2.39 | Original, KWIA, TGV,<br>Train-from-scratch,<br>Fine-tune | 1.77±0.79 | Original, KWIA |

**Fig. 6.**
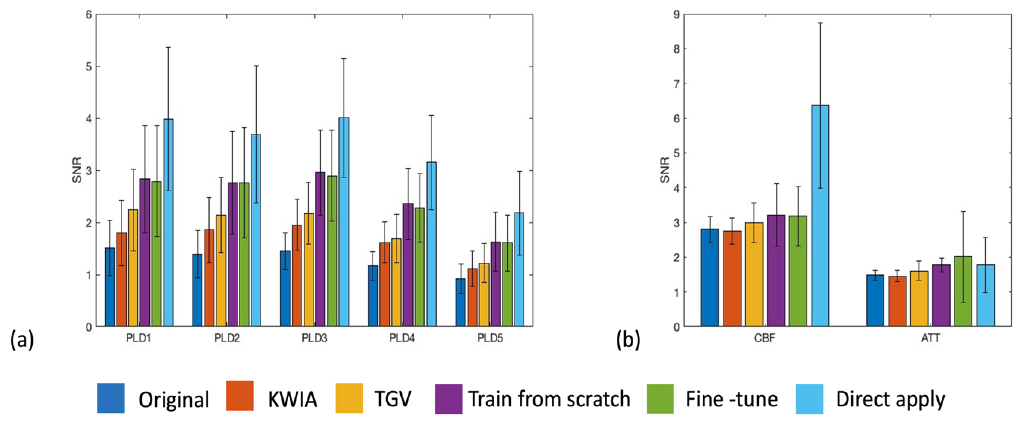
Overall performance of different denoising methods regarding the SNR of perfusion images (a) and in CBF and ATT maps (b).

For perfusion images, KWIA can improve SNR by an average about 30%, which is consistent with the simulation results in a 2-ring KWIA setting[6]. DL trained with KWIA image as reference can improve the SNR of the perfusion images by an average of 92%. For CBF and ATT maps, KWIA does not have much improvement on the performance due to correlated noises between time frames, which is consistent with previous results[6]. But DL model trained with KWIA reference shows an average of 13.5% improved SNR in CBF maps and an average of 28% improved SNR in ATT maps. Direct applying the model can significantly improve the SNR performance but will result in image blurring and bias in quantification values, which will be shown in the following section.

### B. Systematic bias of the quantitative maps

Figure 7 shows CBF and ATT values in whole brain, GM and WM processed with different methods. It shows that KWIA denoising has the smallest difference compared to the original values, which is consistent with the results from previous studies for KWIA denoising. TGV constrained reconstruction will increase CBF values in the whole brain and especially in the WM. Both DL models trained with KWIA denoised images show a small decrease in the CBF values, which is about 10% of the original values. Direct applying the pretrained model shows a large decrease in the CBF values in the WM. For ATT values, TGV reconstructed images show relatively large difference in the whole brain, which means the algorithm does not preserve the temporal relationship across PLDs. KWIA and DL-based methods have relative smaller difference, which shows the consistency of the denoised images with the original images. The details of systematic difference for different methods are shown in Table III.

**TABLE III.** averaged difference of CBF and ATT values in the whole brain, GM and WM areas, mean value and the 95% confidence interval.

| CBF | Whole brain | GM | WM | ATT | Whole brain | GM | WM |
| --- | --- | --- | --- | --- | --- | --- | --- |
| KWIA | -0.71[-0.79, -0.64] | -0.80[-0.90, -0.70] | -0.60[-0.70, -0.50] |  | 0.00[0.00,0.01] | -0.01[-0.01,0.00] | 0.01[0.01,0.02] |
| TGV | 5.72[4.70, 6.74] | 2.67[0.86, 4.48] | 12.37[10.69, 14.04] |  | -0.08[-0.09, -0.07] | -0.05[-0.06, -0.04] | -0.14[-0.17, -0.12] |
| Train-from-scratch | -4.92[-5.45, -4.39] | -5.08[-5.95, -4.20] | -5.76[-6.43, -5.09] |  | -0.02[-0.03, -0.01] | -0.01[-0.02,0.00] | -0.04[-0.06, -0.02] |
| Fine-tune | -4.97[-5.29, -4.65] | -5.22[-5.90, -4.54] | -5.47[-6.08, -4.85] |  | -0.02[-0.03, -0.02] | -0.01[-0.02,0.00] | -0.05[-0.08, -0.02] |
| Direct<br>apply | -7.36[-7.61, -7.11] | -3.59[-4.04, -3.14] | -15.73[-16.46, -15.00] |  | -0.02[-0.03,0.00] | -0.05[-0.06, -0.04] | 0.01[-0.01,0.04] |

**Fig. 7.**
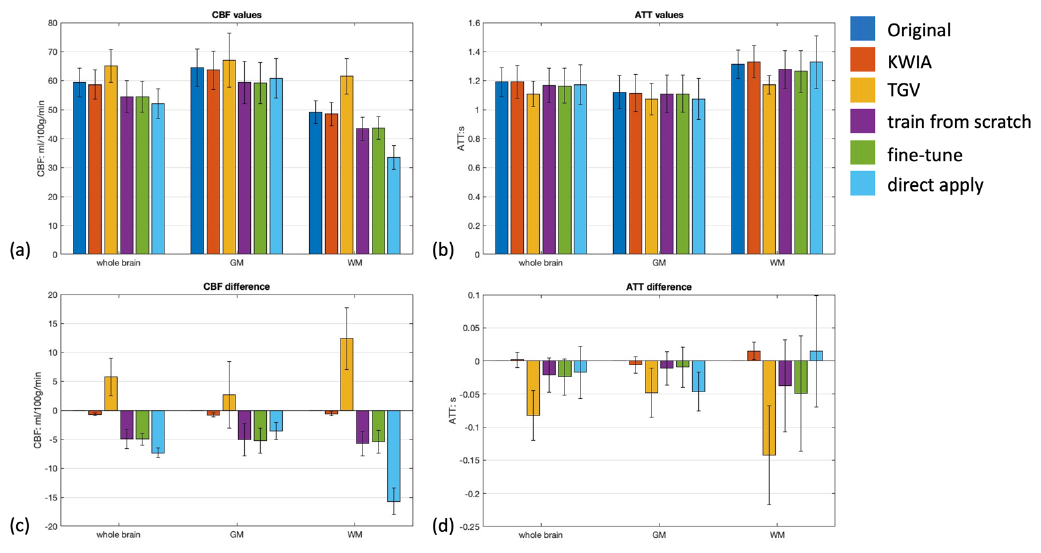
CBF and ATT values processed with different methods (a) and (b) and their relative difference compared to the original values (c) and (d). Error bars show standard deviation.

### C. Test-retest performance on the dataset

Table IV shows the ICC values of whole brain, GM and WM CBF and ATT values processed with different methods. The ICC of the original CBF and ATT images show a moderate repeatability given that the resolution is relatively high for multi-delay ASL, and the reported repeatability in pediatric population is lower than that in adults[20]. TGV-based method has a large decrease in ICC. DL fine-tune method can improve the ICC of whole brain and gray matter CBF and keeps similar ICC values in whole brain, GM and WM ATT values. However, CBF in WM shows a decreased ICC, which indicates the performance of denoising in WM is variable due to the lower SNR and CBF value in the WM.

**TABLE IV.** ICC values of CBF and ATT of test-retest of the 21 subjects with different denoising methods.

| ICC | Original | KWIA | TGV | DL scratch | DL fine tune | DL apply |
| --- | --- | --- | --- | --- | --- | --- |
| Whole brain CBF | 0.40 | 0.38 | 0.05 | 0.42 | **0.45** | 0.47 |
| GM CBF | 0.38 | 0.36 | 0.05 | 0.49 | **0.53** | 0.46 |
| WM CBF | **0.42** | 0.40 | 0.19 | 0.17 | 0.12 | 0.34 |
| Whole brain ATT | 0.59 | 0.60 | 0.58 | 0.62 | **0.63** | 0.58 |
| GM ATT | 0.63 | **0.63** | 0.56 | 0.58 | 0.54 | 0.57 |
| WM ATT | 0.42 | 0.42 | 0.56 | **0.61** | 0.57 | 0.44 |

### D. Comparison of train-from-scratch and fine-tune

To study the difference between training a new model with the new dataset and fine-tune from the pretrained model, we investigated the characteristics of the training process. The performance on the validation dataset across epochs is plotted in Fig. 8. The results are averaged across the five folds. It shows that the fine-tuning method has a higher starting point and faster convergence compared to the train-from-scratch model. But towards convergence, training a new model shows better performance compared to the fine-tuned model, which might indicate the potential overfitting.

**Fig. 8.**
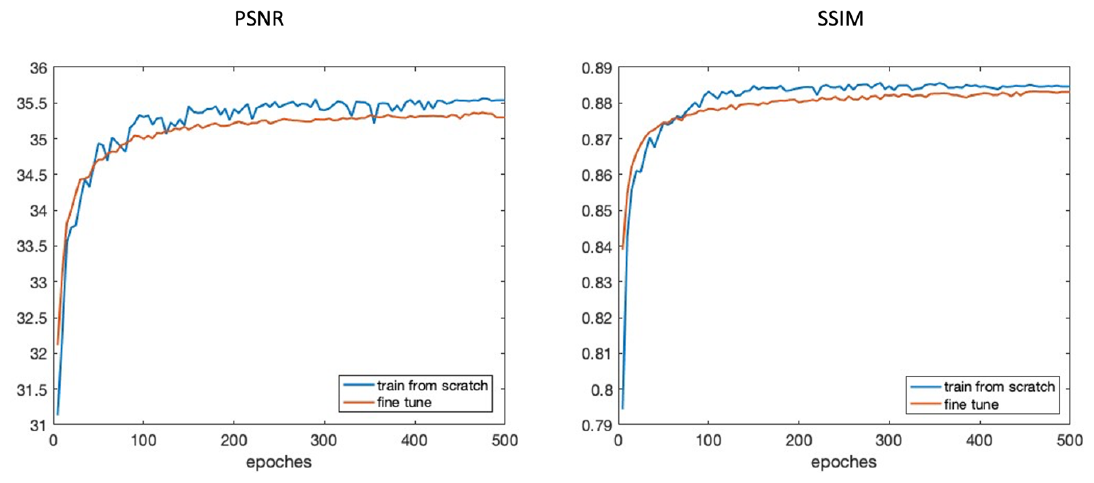
Comparison of training from scratch and fine tune from the pretrained model on their performance on the validation dataset.

## V. DISCUSSION

In this study, we developed a high resolution (iso-2mm) MDASL protocol for pediatric perfusion imaging along with a DL framework with KWIA denoised images as reference for denoising multi-delay ASL. ASL has been shown to be a promising and potentially ideal imaging modality to study neurodevelopment since it is entirely noninvasive and provides perfusion images with higher SNR in children than in adults. Considering the variation of physiological parameters with age (e.g. ATT), MDASL is advantageous for improving perfusion quantification in children. However, it is challenging to achieve high spatial resolution with multi-delay acquisitions due to long acquisition time. The proposed protocol represents a significant step forward compared to the previous study[5] that achieved iso-3mm MDASL with TGV regularized reconstruction, given that the SNR of iso-2mm was ∼30% of that of iso-3mm scan. We proposed a Transformer based DL model to address the low SNR with KWIA denoised images as reference. Our results showed that the proposed DL model outperformed conventional methods such as TGV and KWIA, with high computational efficiency.

There has not been many publicly available ASL datasets especially for multi-delay ASL at a very high-resolution. In previous work for ASL denoising[8], [9], [10], [11], it always requires high-SNR reference images to train the model. Although traditional methods such as Gaussian smoothing or NLM can be applied and serve as a surrogate supervision for training the model, these methods will over smooth the images and are not suitable for the case where the resolution of the image needs to be preserved. Compared to other methods, such as TGV, KWIA has the benefit of easy implementation, minimal spatially blurring and preserving the temporal resolution of the original image series[6]. KWIA uses current time frame and its adjacent time frames to perform weighted averaging in the k-space so that the high frequency part can get more averages and thus higher SNR. Although this will not cause spatial blurring to the image, there will be correlation in the noise among these denoised images, which can cause temporal smoothing to the whole time series. For this reason, it has been shown that little improvement was achieved for the quantitative maps such as CBF and ATT. However, since DL model treats each of the images in the time series independently, the increase in the SNR in all images will result in an overall improvement for the fitting result, which is also shown in our study.

For DL denoising algorithm in medical imaging and especially for quantitative methods, image quality is not the only criteria, the importance of quantification accuracy should also be emphasized. In our experiments, we tried to minimize the quantification biases by using KWIA filtered image as the reference. However, the results still show a small degree of decrease in the fitted CBF maps. Since the value of CBF correlates with the averaged values of the 5 perfusion images, it’s possible that this decrease in value is the result of the removed noise in the perfusion images. ATT values show little changes after denoising, which shows that the temporal relationship among these PLDs has been kept after denoising.

The performance of fine-tuning on a pretrained model is also studied in our experiments. In fine-tuning, the weights of the pretrained model are used as the starting point for the new model instead of a randomized initialization. The fine-tuning strategy is recommended especially when there is not enough training data for the target task since training from scratch might lead to overfitting without enough data. However, it is not clear how to determine if a sample size is small or how will this affect the final results. We tested this by comparing training a new model from scratch and fine-tuning a pretrained model with a similar task, which is single-delay ASL denoising. Compared to the new task, the pretrained model was trained on images with lower resolution and also with the high-SNR reference. The expected SNR improvement for the pretrained model is higher. However, although the task for denoising is similar, when directly applying the pretrained model to denoising the high-resolution multi-delay ASL, the performance is not very good. This is due to the reason that there are still considerable differences between the source and target data, including different resolution, different ASL parameters and different population. In our results, we have shown that fine-tuning from the pretrained model can provide a higher starting point and also faster convergence compared to training a new model from scratch, while the performance of the two models are similar. This would be beneficial when there is only limited training resource and a more generalized model is warranted which fits a larger variety of data.

There are several limitations of this study. First, we do not have a gold-standard reference for our dataset. Our study population is children, and it is not easy to acquire the high-SNR reference for this dataset due to long scan time and head motion. We tested our performance based on non-reference metrics such as SNR. It is warranted that this method to be tested on other datasets with a real reference values. Second, we only used a five-delay dataset. While it is still not clear whether increasing the number of delays will improve the accuracy of quantitative maps, the performance of KWIA improves with the number of rings. With only five delays, we can only use a two-ring KWIA. Nevertheless, a three-ring KWIA is more desirable for datasets with more than 5 delays. Third, whether the quantitative bias reflects a successful denoising remains not clear. Future studies will need to involve larger datasets with ground truth values.

## VI. Conclusion

In this study, we presented a high resolution (iso-2mm) MDASL protocol for pediatric perfusion imaging along with a DL framework with KWIA-denoised image as a surrogate reference for model training, which improves the performance of both perfusion images and the fitted CBF and ATT maps, as well as their test-retest repeatability in the whole brain. This may facilitate future application of multi-delay ASL acquisition in neurodevelopment studies.

## Data Availability

All data produced in the present study are available upon reasonable request to the authors

